# A second dramatic rise in seroprevalence rates of SARS-CoV-2 antibodies among adult healthy blood donors in Jordan; have we achieved herd immunity?

**DOI:** 10.1101/2021.08.15.21261584

**Authors:** Maher A. Sughayer, Asem Mansour, Abeer Al Nuirat, Lina Souan, Rashid Abdel-Razeq, Mahmoud Siag

## Abstract

**Objectives:** To determine the impact of the second wave of COVID-19 and the vaccination campaign on the seroprevalence rates of SARS-CoV-2 antibodies among healthy blood donors in Jordan.

**Methods:** Sera from 536 healthy adult blood donors collected in June -2021 were tested using a commercially available quantitative assay for the total antibodies including IgG against the spike (S) protein receptor binding domain (RBD) of the SARS-CoV-2.

**Results:** 399 (74.4%) of the donors tested positive for the antibodies of whom 69 (17.3%) were confirmed to have been previously infected, 245(61.4%) have received at least one dose of the vaccine and 123(30.8%) were neither diagnosed nor vaccinated. The seropositive donors were significantly more likely to have been vaccinated or previously infected.

**Conclusion:** The crude seroprevalence rate of 74.4% among this group of healthy donors may be encouraging in terms of approaching herd immunity, however with predominance of the delta variant and the uncertainty regarding the required level of herd immunity this goal appears to be far from full achievement in Jordan.

---

In a previous issue of International Journal of Infectious Diseases (Sughayer et al, 2021), we reported a dramatic rise in seroprevalence rates of SARS-CoV-2 in healthy blood donors from 0% during the months of January to September 2020 up to 27.4% in late January-early February 2021, just after the end of the first wave of COVID-19 in Jordan. None of the blood donors had received any vaccination by that time.

A second wave of COVID-19 infection started in March and ended in May, 2021. Also a national vaccination campaign has been ongoing since January of this year. To date approximately 3 million adults have received at least one dose of the COVID-19 vaccine and around 770,000 were PCR-confirmed COVID-19 infected (Ministry of Health-Jordan, 2021). Currently 90% of the new cases are caused by the delta variant.

To assess the impact of the second wave of COVID-19 and the vaccination on the seroprevalence of SARS-CoV-2 we tested 536 healthy blood donors in June -2021 using a commercially available quantitative assay for the total antibodies including IgG against the spike (S) protein receptor binding domain (RBD) of the SARS-CoV-2: the Elecsys Anti-SARS-CoV-2 S (Roche Diagnostics, Mannheim, Germany). This assay uses a recombinant protein representing the RBD of the S antigen in a double-antigen sandwich assay format, which favors detection of high affinity antibodies against SARS⍰CoV⍰2. It has a sensitivity of 98.8 % (95 % CI: 98.1 – 99.3 %) and a specificity of 100 % (95 % CI: 99.7 – 100 %).

The demographics of the blood donors and the results of the serological assay are shown in the table.

| Category |  | Seropositive donors<br>399 of 536 | Seronegative donors<br>137 of 536 | p-value | Crud Prevalence rate |
| --- | --- | --- | --- | --- | --- |
| Number of donors |  | 399 (%) | 137 (%) |  | 74.4% |
| Male |  | 361 (90.5%) | 119 (86.9%) | 0.23 | 75.2% |
| Female |  | 38 (9.5%) | 18 (13.1%) |  | 67.9% |
| Age (yr) | 18-30 | 204 (51.1%) | 85(62.1%) | 0.06 | 70.6% |
|  | 31-40 | 115(28.8%) | 29(21.2%) |  | 79.9% |
|  | 41-50 | 52(13.0%) | 19(13.9%) |  | 73.2% |
|  | 51-68 | 28(7.1%) | 4(2.9%) |  | 87.5 % |
| Blood group & RH: | O | 143(35.8%) | 50(36.5%) | 0.12 | 74.1% |
|  | A | 145(36.3%) | 62(45.3%) |  | 70.0% |
|  | B | 84(21.1%) | 18(13.1%) |  | 82.4% |
|  | AB | 27(6.8%) | 7(5.1%) |  | 79.4% |
| RH positive |  | 351(88.0%) | 122(89.1%) | 0.73 | 74.2% |
| RH Negative |  | 48(12.0%) | 15(10.9%) |  | 76.2% |
| Residential Location North |  | 67(16.8%) | 26(18.9%) | 0.48 | 72.0% |
| Center including Amman |  | 319(79.9%) | 109(79.6%) |  | 74.5% |
| South |  | 13(3.3%) | 2(1.5%) |  | 86.7% |
| Covid-19 | Confirmed Past infection | 69(17.3%) | 2(1.5%) | <0.00001 |  |
|  | Vaccinated | 245(61.4%) | 33(24.1%) | <0.00001 |  |
|  | No/negative PCR | 330(82.7%) | 135(98.5%) | <0.00001 |  |
|  | No infection/No vaccine | 123(30.8%) | 103(75.2%) | <0.00001 |  |
|  | Confirmed past infection/No vaccine | 31 (7.8%) | 1 (0.7%) | 0.0027 |  |

As seen in the table 399 (74.4%) of the donors were found to be seropositive of whom 245 (61.4%) have received at least one dose of the vaccine and 69 (17.3%) were previously infected. Some of the previously infected have received also at least one dose of the vaccine. A substantial number 123 (30.8%) of the seropositive donors were neither vaccinated nor had been previously diagnosed with the infection. Interestingly, 33 (24.1%) of the seronegative donors have received the vaccine of whom 29 received only one dose and 4 received 2 doses. While of the vaccinated seropositive donors 119 have received only one dose of the vaccine in contrast to 126 who received full vaccination.

The vaccines types were either Pfizer-BioNTech, Sinopharm, or Astra-Zeneca’s. No significant differences were found between the seronegative and seropositive donors in terms of age, sex, blood group or residential location. However the seropositive donors were significantly more likely to have been vaccinated or previously infected. The overall at least one dose-vaccination rate in the entire group of donors is 51.9%. The one-dose vaccination rate in the targeted population of Jordan is currently approximately 45%.

The crude seroprevalence rate of 74.4% in this group of blood donors may indicate that the Jordanian adult community has attained a substantial rate of herd immunity. The most important question then becomes: what is the level of herd immunity needed in the case of COVID-19 to effectively prevent the transmission?

The minimum threshold for herd immunity is theoretically estimated to be 60-72%, however the issue is much more complicated and it may be up to 90% and even that the entire population needs to be vaccinated/revaccinated to cut the transmission (Kadkhoda, 2021; Aschwanden, 2020). Therefore we believe that despite of this apparent high level of community immunity we are far from achieving the required herd immunity. Continuation of the vaccination campaign and other precautionary measures of mask-wearing and social distancing are still need to be adhered to.

## Data Availability

Data is available upon request

The Authors declare no conflict of interest.

No funding was received.

The IRB at King Hussein Cancer Center approved the study

